# Task-Specific Quality Gating for Retinal Optical Coherence Tomography B-Scans: Learned Representations Over Scalar Metrics in Choroid Segmentation

**DOI:** 10.64898/2026.09.14.26363079

**Authors:** Amritesh Hiras, Aditya Jayaraman, Adarsh Gadari, Anuj Sharad Mankumare, Akshith Mynampati, Jay Chhablani, Sandeep Chandra Bollepalli, Anand Kakarla, Kiran Kumar Vupparaboina

## Abstract

Automated segmentation of Optical Coherence Tomography (OCT) images is a critical component of structural biomarker extraction for retinal diagnostics. Deep learning models achieve state-of-the-art performance on controlled datasets, yet exhibit unpredictable failures on real-world data. Current quality gates rely on device-reported scan quality scores, which have been shown to be unreliable predictors of segmentation performance. We define scan quality in a task-specific sense that is, whether a given B-scan will yield a reliable segmentation from a particular trained model. Under this definition, we perform a systematic evaluation of No-Reference Image Quality Assessment (NR-IQA) metrics, general-purpose and domain-specific pretrained representations as alternative quality gates. To this end, we use choroid segmentation as the prototype task, with a dataset of 6,076 OCT B-scans from 80 subjects. These quality gate candidates are evaluated at three levels: scalar metrics (BRISQUE, NIQE, PIQE, SNR, PSNR), supervised linear probing, and unsupervised partitioning (K-Means) of the feature vectors and the learned representations. All scalar NR-IQA metrics proved inadequate (|*r*| *<* 0.20). General-purpose ImageNet-based pretrained representations (EfficientNet-b0, ResNet-50, ViT-B/16) improve upon NR-IQA, achieving ROC-AUC up to 0.77, indicating that learned representations are better suited to task-specific quality gating than hand-crafted scalar statistics. Retinal foundation models (FMs) further close the gap: RETFound (OCT-specific FM) achieves ROC-AUC ≈ 0.81. Unsupervised K-Means partitioning indicates that general-purpose ImageNet-pretrained embeddings, despite carrying a linearly decodable quality signal, do not reliably organize scans by quality geometrically, whereas the retinal FMs produce quality-aligned clusters that exceed a patient-level permutation null, suggesting that domain-specific pretraining provides additional, complementary benefit on top of general-purpose learned representations.

## I. Introduction

Automated segmentation in Optical Coherence Tomography (OCT) is increasingly embedded in clinical pipelines, where deep learning models achieve strong performance on controlled datasets [1], [2] but degrade unpredictably on real-world scans [3], [4], motivating mechanisms that flag unreliable model outputs before they propagate through downstream workflows [5]. Importantly, whether an acquired scan is adequate depends not only on its imaging quality but also on the downstream task and model: a scan that is adequate for one task/model may have insufficient information for another. A reliable deployment therefore requires a task-specific, pre-inference quality gate that determines whether a deployed model is likely to produce a trustworthy output.

Prior work on predicting reliability has largely focused on output-based uncertainty quantification [6] or input-based out-of-distribution detection [7], asking whether the model is uncertain or the input is distributionally novel. We instead assess task suitability of in-distribution inputs: scans consistent with the model’s expected distribution that nevertheless yield unreliable predictions. The prevailing clinical quality gate, the device-reported signal-strength index (e.g., Heidelberg Q-Score), is a poor proxy for this — Gadari et al. showed it explains less than 1.4% of segmentation variance (*R*^2^ *<* 0.014) across 5,047 B-scans [8]. No-Reference Image Quality Assessment (NR-IQA) methods such as BRISQUE [9], NIQE [10], and PIQE [11] estimate quality from natural scene statistics without a reference image, but their ability to predict downstream segmentation reliability in OCT remains untested. Recent OCT-specific frameworks such as AQUA-OCT and ROQUS [12], [13] similarly assess perceptual quality but do not evaluate whether a specific segmentation model will succeed on a given scan — motivating a direct evaluation of image-quality measures for downstream task failure prediction.

We investigate this problem using automated choroid segmentation as a prototype application with a single deployed model, benchmarking 6,076 OCT B-scans from 80 subjects to determine whether existing image-quality measures and learned image representations contain information that can be leveraged to identify segmentation failure pre-inference. We first assess the applicability of scalar NR-IQA measures, then evaluate a hierarchy of representations — NR-IQA (BRISQUE, NIQE) feature vectors, general-purpose ImageNet-pretrained embeddings (ViT-B/16, ResNet-50, EfficientNet-B0), and retinal foundation models (RET-Found [14], UrFound [15]), using supervised linear probing and unsupervised K-means clustering.

Our results reveal a clear hierarchy: scalar NR-IQA metrics carry little useful signal (|*r*| *<* 0.20) towards identifying segmentation failure and their feature spaces show no quality-discriminative structure. General-purpose ImageNet-pretrained representations improve upon NR-IQA (ROC-AUC ≤ 0.685), achieving an ROC-AUC scores up to 0.773, indicating that learned visual representations capture information relevant to segmentation reliability even without task-specific training. Retinal foundation models surpassed both NR-IQA and general-purpose pretrained representations, with RETFound achieving an ROC-AUC of 0.809. These findings support learned representations as candidate signals for task-specific, pre-inference quality gating, motivating further evaluation across segmentation tasks, models, and datasets.

## II. Related Work

Early OCT segmentation relied on image processing, graph-theoretic methods, active contours, and curvelet transforms [16], [17] before U-Net [2] and subsequent attention- and Transformer-based architectures [18], [19] compete with near-human performance [1]. However, the deployed models still tend to fail unpredictably on individual scans [3], [4].

Quality assessment in OCT has historically mirrored other modalities such as Magnetic Resonance Imaging (MRI) and Computed Tomography (CT), sharing their reliance on global physical parameters such as signal-to-noise ratio [20], operationalized clinically via device-reported indices like the Heidelberg Q-score [8]. The OSCAR-IB consensus criteria [21] and its automated OSCAR-AI extensions formalized the evaluation across physical and algorithmic failure, and more recent frameworks such as AQUA-OCT [12] and ROQUS [13] automate the classification against clinical acquisition guidelines and clinician readability. All of these, however, treat quality as an intrinsic, human-perceived property, leaving whether visual usability reliably translates to deep learning segmentation stability.

The limitations of task-agnostic quality measures have motivated the development of task-specific quality assessment. These studies aim to address this gap directly by evaluating quality through the performance of the downstream model rather than visual readability [22], alongside parallel efforts in ground-truth-free failure detection via error regression [23], reverse classification accuracy [24], and confidence aggregation [25]; clinical OCT studies confirm that scans that meet the manufacturer quality thresholds frequently still suffer catastrophic segmentation failures [21], [26].

## III. Dataset, Quality Labels and Methods

### A. Dataset

This is a retrospective study performed under the tenets of the Declaration of Helsinki with the approval of the Institutional Review Board of the University of Pittsburgh Medical Center. Informed consent of the subject was for the retrospective study. We analyzed 124 enhanced-depth imaging (EDI) OCT volumes from 80 subjects, including 36 subjects with unilateral scans and 44 subjects with bilateral scans. All volumes were acquired using foveal-centered 6 *×* 6 mm^2^ scanning protocols, with each volume comprising 49 B-scans. Ground-truth (GT) choroid boundaries were obtained through manual correction of the automatically generated boundaries for all the 6,076 scans, following an established practice in medical image segmentation [27]. The corrected boundaries were used as the reference standard for evaluating segmentation performance. Automatic choroid segmentations were generated using a Residual U-Net model (NMI-ChoroidAI) trained on a separate dataset of OCT B-scans with choroid annotations [28]. None of the B-scans included in the present study were used to train or fine-tune the model.

Segmentation reliability was quantified using the Dice similarity coefficient (DSC) between the model-generated segmentation and the corresponding GT. Scans yielding DSC below 0.85 were labelled *Bad* (*n* = 2,921, 48.1%) and those at or above as *Good* (*n* = 3,155, 51.9%). Dice scores ranged from 0.29 to 0.98, with a mean of 0.844 *±* 0.076. The threshold was used solely to operationalize segmentation failure for the benchmarking analysis and does not represent an intrinsic boundary of image quality; the proposed framework can be evaluated using alternative thresholds (see, section VI).

### B. Metric-Based Baselines

Five metrics were selected for evaluation: two standard signal-strength measures (SNR and PSNR) alongside three widely used NR-IQA metrics grounded in Natural Scene Statistics (NSS): BRISQUE [9], PIQE [11], and NIQE [10]. We have also extracted feature vectors from BRISQUE and NIQE. Both SNR and PSNR were computed using a Gaussian filter to obtain a denoised reference 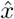:

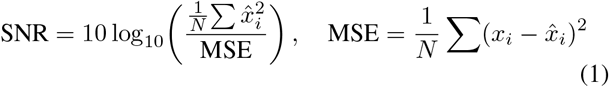

### C. Pre-Trained Models and Probing Protocol

#### a. Frozen embeddings

Embeddings were extracted from five frozen architectures: three general-purpose ImageNet-pretrained backbones — ResNet-50 (2,048-dim), EfficientNet-B0 (1,280-dim), and ViT-B/16 (768-dim) — and two retinal foundation models: RETFound [14], a ViT-Large/16 model (1,024-dim) pretrained on OCT images, and UrFound [15], a multimodal ViT-Base/16 model (768-dim) pretrained on both Color Fundus Photography and OCT images. For the CNNs, the embedding is the network’s native global-average-pooled feature; for all transformer-based models, the embedding is the mean over patch tokens.

#### b) Linear probing evaluation with LOPO

We evaluated each frozen embedding with a linear probe under leave-one-patient-out (LOPO) cross-validation: a logistic-regression probe was trained on all but one of the 80 subjects and evaluated on the held-out subject, repeated so that each subject served as the test fold exactly once. The probe was a single linear layer trained by full-batch L-BFGS on binary cross-entropy with an *ℓ*_2_ penalty; its two hyperparameters — weight decay and an optional class-balanced positive-class weight (pos weight = *n*_neg_*/n*_pos_) — were tuned per fold via nested 3-fold patient-grouped cross-validation on that fold’s training subjects only, using 20 Optuna trials (TPE sampler) with mean inner-fold ROC-AUC as the objective. The tuned probe was refit on the full training set and used once to predict the held-out subject’s images. Predictions from all 80 folds were pooled into a single set of out-of-fold predictions before computing summary metrics; we report pooled F1 and ROC-AUC with 95% confidence intervals from patient-level bootstrap resampling (2,000 resamples with replacement, 2.5th/97.5th percentile endpoints).

#### c) Unsupervised quality separation (K-Means)

We additionally probed each frozen embedding’s geometric structure with K-Means clustering, which imposes no supervised decision boundary. To avoid clustering on patient identity rather than image quality, we resampled the data 300 times, each time selecting one random frame per patient (*n* = 80) and standardizing that resampled set. For each embedding space, the number of clusters *k*^∗^ was selected once, via the Kneedle method (*k* = 1–10) on the within-cluster sum-of-squares curve of the full, non-subsampled standardized embeddings, and held fixed across all 300 resamples. For each resampled clustering we computed the Max-Bias, Min-Bias and the spread: the highest, lowest percentage of Bad-quality scans (DSC *<* 0.85) across clusters and their difference, summarized across the 300 resamples as its median with a 95% interval.

## IV. No-Reference Image Quality Assessment

### A. Scalar Metric Correlations

Table I reports Pearson correlations between each scalar quality metric and per-scan DSC on the full 6,076-image dataset. Standard NR-IQA metrics proved ineffective: BRISQUE yielded *r* = −0.141, PIQE *r* = −0.081, and NIQE *r* = −0.030, none of which offer actionable signal. Signal strength based metrics follows NSS metrics, with SNR at *r* = −0.194 and PSNR at *r* = −0.072. Across all five metrics, the highest observed correlation is |*r*| = 0.194 (SNR), which is insufficient for reliable automated quality gating. This result suggests that full image metrics aggregated into a scalar metric does not provide sufficient signal for task-specific quality gating, specifically in AI-based pipelines.

**TABLE I.** Pearson Correlation of Quality Metrics with Segmentation DSC.

| Metric | Type | Pearson $r$ |
| --- | --- | --- |
| BRISQUE | NR-IQA (NSS) | −0.141 |
| NIQE | NR-IQA (NSS) | −0.030 |
| PIQE | NR-IQA (NSS) | −0.081 |
| SNR | Signal Strength | −0.194 |
| PSNR | Signal Strength | −0.072 |

### B. Linear Probing NR-IQA Feature Vectors

Table II shows that pooled ROC-AUC for the pre-aggregation NR-IQA feature vectors were better than chance: BRISQUE reached 0.685 [0.618, 0.743] and NIQE 0.674 [0.607, 0.735]. F1 was comparable across two metrics (0.617/0.620 for BRISQUE/NIQE), reflecting the class-balanced operating point rather than discriminative ability. These results indicates that BRISQUE and NIQE’s failure as task-specific scalar quality scores lies in the aggregation step: their underlying 36-dimensional NSS feature vectors carry quality-relevant information that a linear probe recovers, even though the scalar score built from them does not.

**TABLE II.** Linear probe performance for Good (DSC ≥ 0.85) versus Bad (DSC *<* 0.85) quality classification, evaluated via leave-one-patient-out cross-validation. Pooled test F1 and ROC-AUC are reported as point estimate [95% CI], with confidence intervals from patient-level bootstrap resampling.

| Model | F1 | ROC-AUC |
| --- | --- | --- |
| BRISQUE | 0.617 [0.528, 0.695] | 0.685 [0.618, 0.743] |
| NIQE | 0.620 [0.531, 0.702] | 0.674 [0.607, 0.735] |
| ResNet-50 | 0.671 [0.590, 0.740] | 0.748 [0.693, 0.797] |
| EfficientNet-B0 | 0.680 [0.605, 0.744] | 0.744 [0.688, 0.794] |
| ViT-B/16 | 0.700 [0.623, 0.765] | 0.773 [0.722, 0.820] |
| UrFound | **0.729** [0.663, 0.784] | 0.803 [0.757, 0.845] |
| RETFound | **0.729** [0.664, 0.785] | **0.809** [0.764, 0.849] |

### C. Unsupervised Linear Partitioning of NR-IQA feature vectors

The optimal number of clusters was determined by the elbow method described in Section III-C (BRISQUE: *k*^∗^ = 4; NIQE: *k*^∗^ = 3;). None of the two NR-IQA feature spaces separated Bad-from Good-quality scans beyond chance under the permutation test (Table III): BRISQUE’s Min-Bias/Max-Bias spread of 46.9 percentage points did not exceed its permutation null of 33.4 (*p* = .426); NIQE’s spread of 26.3 was likewise indistinguishable from its null of 16.5 (*p* = .281). In each case, a spread of similar magnitude arises from the max-minus-min selection over *k*^∗^ noisy cluster rates even when the Bad/Good labels are randomly shuffled, so none of these nominal contrasts constitute evidence that the NR-IQA embedding geometry organizes scans by segmentation quality.

**TABLE III.** Unsupervised K-Means quality separation at the patient level across 300 resamples. Results are summarized as cluster bias and Spread (Max-Bias−Min-Bias); ^∗^*p <* .05, ^∗∗^*p <* .01.

| Model ($k^*$) | Min-Bias | Max-Bias | Spread [95%] | Null [95%] |
| --- | --- | --- | --- | --- |
| BRISQUE (4) | 36.8 | 71.4 | 46.9 [9.2, 72.7] | 33.4 [7.1, 70.6] |
| NIQE (3) | 37.5 | 65.5 | 26.3 [6.7, 52.8] | 16.5 [3.0, 43.4] |
| ResNet-50 (5) | 0.0 | 100.0 | 78.7 [44.4, 100.0] | 71.4 [29.2, 100.0] |
| EfficientNet-b0 (3) | 39.3 | 57.1 | 19.1 [3.1, 55.6] | 17.9 [3.3, 55.0] |
| ViT-B/16 (4) | 16.7 | 66.7 | 49.0 [22.0, 70.8] | 23.2 [7.1, 47.5] |
| RETFound* (3) | 17.4 | 65.6 | 47.1 [31.2, 65.1] | 15.6 [2.9, 36.2] |
| UrFound** (3) | 17.4 | 66.7 | 48.9 [32.1, 68.8] | 15.8 [3.0, 36.3] |

ResNet-50 has unreliable estimates.

## V. Pretrained Embeddings and Foundation Models

### A. Linear Probe on Frozen Embeddings

Table II reports pooled leave-one-patient-out (LOPO) linear-probe ROC-AUC and F1 across all representations. Among ImageNet-pretrained backbones, ViT-B/16 achieves the highest pooled ROC-AUC (0.773). A paired patient-level bootstrap test (5,000 resamples) confirms this margin is reliably non-zero relative to both ResNet-50 (ΔROC-AUC = 0.025, *p* = 0.004) and EfficientNet-B0 (ΔROC-AUC = 0.029, *p* = 0.002), though the margin itself is modest. ResNet-50 and EfficientNet-B0 are statistically indistinguishable from each other (ΔROC-AUC = 0.004, *p* = 0.63). As a group, the ImageNet-pretrained backbones exceed the NR-IQA baselines (BRISQUE 0.685, NIQE 0.674) by a substantially larger and equally reliable margin (ΔROC-AUC = 0.076, *p <* 0.0001).

Retinal foundation models extend this ranking further. A paired patient-level bootstrap test confirms the foundation-model group (UrFound, RETFound) exceeds the ImageNet-pretrained group (ResNet-50, EfficientNet-B0, ViT-B/16) as a whole (ΔROC-AUC = 0.051, 95% CI [0.023, 0.080], *p <* 0.001), and both groups significantly exceed the NR-IQA baselines (ImageNet-pretrained: ΔROC-AUC = 0.076; foundation models: ΔROC-AUC = 0.127; *p <* 0.0001 in both cases). Critically, the same test finds no statistically significant difference between UrFound and RETFound themselves (ΔROC-AUC = 0.006, 95% CI [− 0.008, 0.021], *p* = 0.43), despite UrFound’s multimodal, text-supervised pretraining and RETFound’s OCT-only unimodal pretraining – suggesting that domain relevance of the pretraining data, rather than the specific supervision recipe within that domain, is the primary factor separating these two groups.

### B. Geometric Structure of Embedding Spaces

K-Means was applied to all five frozen pretrained architectures, with *k*^∗^ determined via elbow analysis and cluster quality-enrichment tested against a patient-level permutation null (Table III). ResNet-50 (*k*^∗^ = 5) and EfficientNet-b0 (*k*^∗^ = 3) did not exceed their permutation nulls (*p* = .574 and *p* = .490); ResNet-50’s nominal Min-Bias/Max-Bias extremes (0.0%/100.0% Bad) reflect an unstable partition rather than a genuine effect. ViT-B/16 (*k*^∗^ = 4) showed a larger spread than either CNN (49.0 vs. its null of 23.2) but fell short of significance at this sample size (*p* = .073). The two retinal foundation models surpassed the permutation null: UrFound (*k*^∗^ = 3) partitioned into a Min-Bias cluster of 17.4% Bad and a Max-Bias cluster of 66.7% Bad, a spread of 48.9 against a null of 15.8 (*p* = .009), and RETFound (*k*^∗^ = 3) showed an almost identical partition – Min-Bias 17.4%, Max-Bias 65.6% Bad, spread 47.1 against a null of 15.6 (*p* = .010). Only for these two representations does the observed spread exceed what label-permutation alone produces by a wide margin, indicating that OCT-specific pretraining makes segmentation quality a geometrically salient, unsupervised axis of the representation.

## VI. Discussion

The better performance of learned image representations compared with conventional NR-IQA representations suggests that information relevant to segmentation reliability is encoded in the learned visual feature space. General-purpose ImageNet-pretrained representations outperformed both scalar NR-IQA measures and NR-IQA feature vectors, despite having no exposure to OCT images or choroid segmentation during pretraining (Table II). Importantly, this does not imply that ImageNet representations are inherently optimized for quality assessment; instead, their performance demonstrates that representations learned for a different visual recognition objective can contain transferable information relevant to identifying segmentation failure.

To separate the contribution of the learned weights from that of the network architecture itself, we repeated the linear-probing analysis using features from randomly initialized (8 different seeds), untrained backbones of the same architectures. Random-initialization features still exceeded the scalar NR-IQA metrics (pooled LOPO ROC-AUC ≈ 0.70–0.71 for all five architectures, versus 0.68 and 0.67 for BRISQUE and NIQE), indicating that part of this signal reflects generic architectural inductive biases rather than anything learned during pretraining [29]. Pretraining nonetheless added a consistent increment on top of this baseline, and this increment was roughly two to three times larger for the retinal foundation models (≈ +0.10 ROC-AUC) than for the ImageNet backbones (≈ +0.03–0.07); because the randomly initialized foundation-model architectures performed no better than the randomly initialized ImageNet architectures (all ≈ 0.70– 0.71), the advantage of RETFound and UrFound is plausibly attributable to their domain-specific pretraining rather than to their architecture. This pattern was not an artifact of the specific Dice threshold (0.85) used to define good quality segmentation: sweeping the threshold from 0.80 to 0.95 (Fig. 3), the difference between pretrained and random initialization for the foundation models remained positive and narrowed with increasing threshold. The same pattern is observed for ImageNet backbones and reversed sign from 0.92 onward for two of the three backbones (ResNet-50, ViT-B/16), where roughly 15% of scans are still labelled good quality. At the most stringent thresholds (≥ 0.94, fewer than 10% of scans labelled good quality) do all three ImageNet backbones’ estimates become comparably unstable, while RETFound and UrFound diminished their advantage after 0.94. The benefit of domain-specific pretraining is thus robust to the choice of operating point, however, may be useful in identifying truly bad images as their advantage is predominantly seen towards lower threshold.

**Fig. 1.**
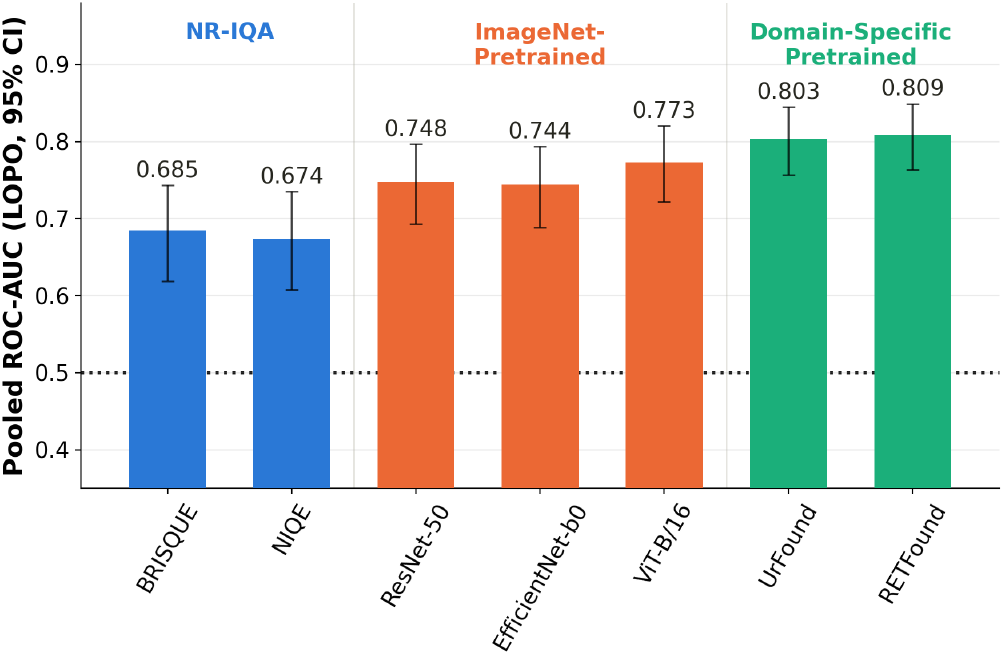
Choroid-segmentation quality-gating performance by feature source: no-reference image-quality (NR-IQA) feature vectors (BRISQUE, NIQE), ImageNet-pretrained backbones (ResNet-50, EfficientNet-B0, ViTB/16), and domain-specific foundation models pretrained on retinal imaging (UrFound, RETFound). Figure shows pooled ROC-AUC under the leave-one-patient-out (LOPO) protocol (n=80 patients) with error bars showing the 95% patient-level bootstrap confidence interval. The dotted line marks chance-level discrimination (AUC = 0.5). NR-IQA metrics show modest discriminative value, ImageNet pretraining offers a limited improvement, and domain-matched foundation-model pretraining performs better than both groups.

**Fig. 2.**
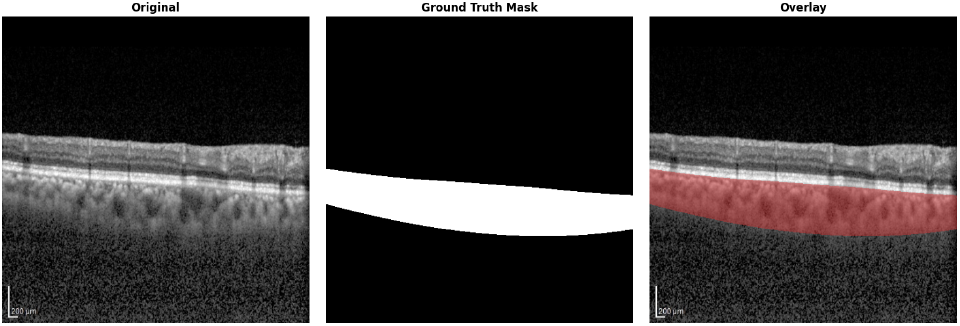
Representative OCT B-scan (grayscale) with the automatically predicted choroid segmentation boundary overlaid (coloured contour). The choroid lies between the retinal pigment epithelium and the sclera; accurate delineation is required for reliable thickness measurements. Image from the 124-volume, 103-eye dataset used in this study.

**Fig. 3.**
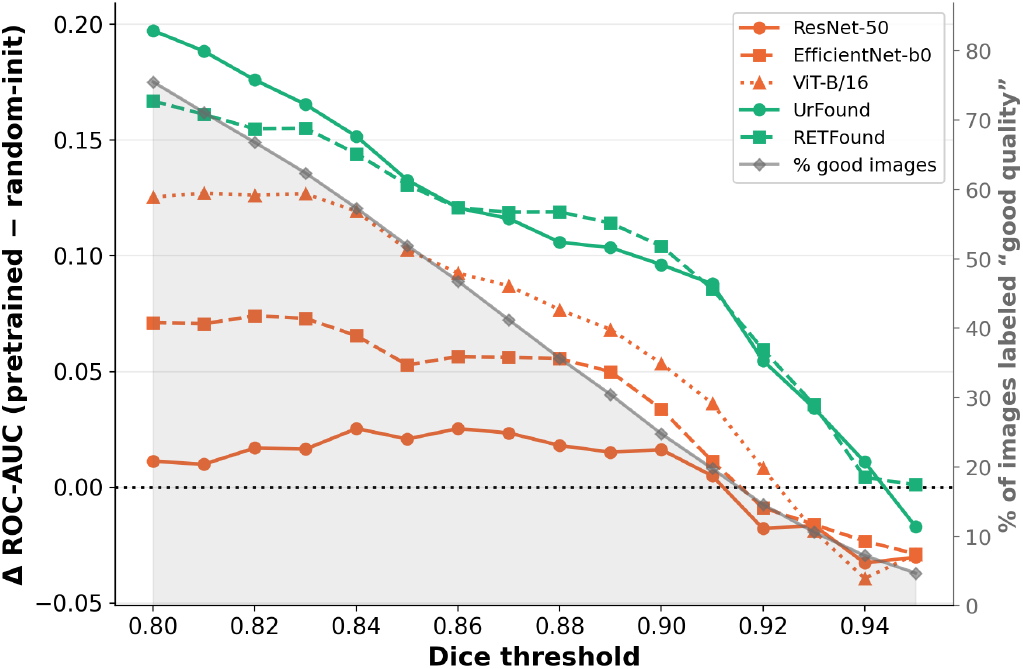
Pooled LOPO ΔROC-AUC (pretrained v random-init) versus the Dice threshold defining good segmentation, across 0.80–0.95. Grey shading (right axis) shows the corresponding class-balance shift – % of the images labeled Good at each threshold. Domain-matched models retain a positive pretraining advantage, although decreasing towards the higher threshold; ImageNet backbones’ advantage narrows and reverses at the most stringent thresholds, where the Good class becomes a small minority.

With cluster count fixed at *k* = 2 to match the true binary quality label, pretraining had opposite effects depending on the pretraining domain (Fig. 4). For the generic ImageNet-pretrained backbones, pretraining significantly reduced how well blind 2-way clustering recovered the quality split relative to random initialization (ResNet-50: ΔSpread = − 19.7 pts; ViT-B/16: −24.9 pts; both *p <* 10^−30^), while EfficientNet-B0 showed no reliable difference at this threshold (ΔSpread = 0.1 pts, *p* = 0.58). The retinal foundation models showed the opposite pattern: pretraining significantly increased the quality-aligned spread relative to random initialization (UrFound: +9.7 pts; RETFound: +6.6 pts; both *p <* 10^−25^), and this advantage held across most of the threshold range before narrowing and reversing at the most stringent thresholds (≥ 0.94), the same small-positive-class regime where the supervised ROC-AUC results also became unstable. Generic ImageNet pretraining thus appears to actively reorganize the embedding space away from quality-relevant structure, whereas domain-specific pretraining reorganizes it toward quality-relevant structure – a geometric counterpart to the ROC-AUC findings above.

**Fig. 4.**
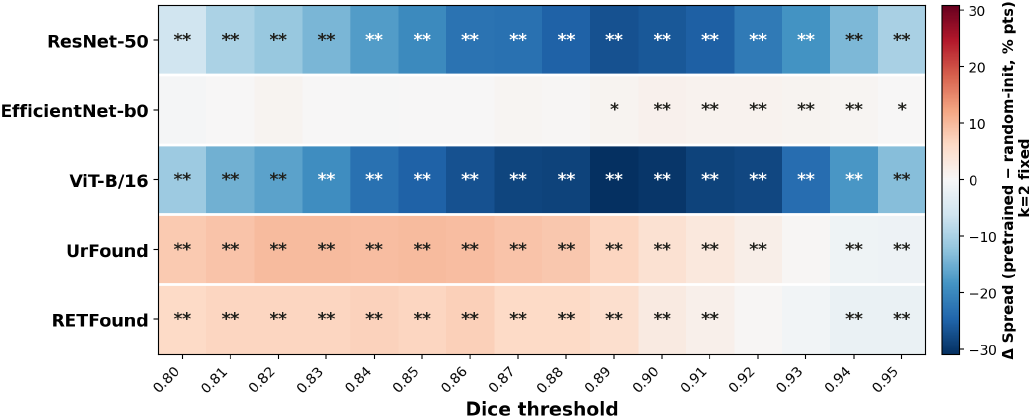
Paired difference in unsupervised quality separation (pretrained random-init) across Dice thresholds, with *k* = 2 fixed to match the binary Good/Bad labels. Values show the median difference in Spread across 300 matched patient-level resamples; red/blue indicate increased/decreased separation with pretraining. Asterisks denote paired Wilcoxon signed-rank significance (^∗∗^*p <* 0.01, ^∗^*p <* 0.05). At the most stringent thresholds (≥ 0.94), fewer than 10% of scans are labeled Good.

### A. Clinical Implications

A reliable choroid quality gate does not require bespoke architecture design: a frozen retinal foundation model backbone with a supervised linear head trained on DSC already achieves ROC-AUC of 0.809, while being efficient in identifying scans at lower threshold. The critical design requirements are that quality labels be task-specific — derived from segmentation model performance rather than human readability — and that the backbone be pretrained on the target modality. We believe, such a system can reduce the image rejection rates [30].

### B. Limitations and Future Work

This study is limited to a single OCT device and healthy patient population; future work will extend evaluation across multiple devices and diverse pathology cohorts. The study is limited to evaluating pretraining data rather than the training objective, this nuance is left for the future study. We have not established the causal signal for these results rather the results are encouraging and warrants further research. More generally, task-specific quality gating based on learned representations warrants validation against additional image attributes, including features such as choroidal boundary visibility that are not explicitly evaluated here. Finally, given the few-shot capabilities of foundation models, evaluating their use for quality gating with limited task-specific data represents an important direction for future work.

## VII. Conclusion

This study evaluated existing image-quality measures and learned image representations as candidate signals to identify pre-inference segmentation failure in OCT choroid segmentation. Conventional NR-IQA measures provided limited information about downstream segmentation reliability, whereas learned visual representations contained stronger signals. The progressive improvement from general-purpose pretrained representations to retinal foundation models, further suggests that domain-relevant pretraining can provide representations better suited to task-specific quality assessment. Together, these findings support a shift from generic image-quality assessment toward task-specific quality gating, in which the suitability of an acquired scan is evaluated in relation to its intended downstream analysis. Although demonstrated here as a prototype for choroid segmentation, this framework provides a basis for evaluating whether learned representations can support pre-inference quality assessment across other medical imaging tasks. Further validation across datasets, tasks, and segmentation models is needed to establish its generalizability and clinical utility

## Data Availability

Data is currently not available publicly, however, will be reviewed upon request case-by-case basis.

